# Community-associated Carbapenem-Resistant Organism Case Investigations in New York City

**DOI:** 10.1101/2025.03.14.25323914

**Authors:** Celina Santiago, Rebecca Zimba, Ying Lin, Ulrike Siemetzki-Kapoor, Nicole Burton, Katelynn Devinney, Dominique Balan, Thomas Portier, William G. Greendyke, Molly M. Kratz, Kailee Cummings, Catharine Prussing, Faten Taki, Saymon Akther, Karen A. Alroy

## Abstract

**Background:** Community-associated carbapenem-resistant organisms (CA-CRO) are a growing concern. The New York City (NYC) Health Department sought to identify, quantify, and characterize CA-CRO in NYC.

**Methods:** CA-CRO cases were gram-negative carbapenem-resistant bacteria, cultured from urine or skin, collected December 2020–May 2023 among NYC residents aged ≤70 years with no international travel, hospitalizations, or long-term care facility stays within 12 months before specimen collection. Data were from laboratory-based surveillance, medical records, and patient interviews asking about medical and behavioral history. Sequencing was conducted to explore potential genomic clustering.

**Results:** Among 114 patients eligible after chart review, 75 were reached for screening. Of those, 36 met the case definition and were interviewed: 61% were female; 39% Latino, and 19% Black; median age was 61 years; and 36% lived in high/very high poverty areas. Fifty-eight percent reported ≥1 comorbidity; 35% reported taking antibiotics within 3 months of specimen collection; and 25% had a urinary catheter or indwelling device within 2 days of specimen collection. Only 6 of 15 sequenced isolates clustered with other sequences from public repositories or laboratory databases.

**Conclusions:** CA-CRO were rare. Patients with a CA-CRO were disproportionately female, non-white, and medically complex. Interviews enhanced eligibility screening and facilitated gathering rich medical and behavioral histories. Despite limited sequencing, the preponderance of non-clustering isolates suggested that coverage of CRO sequences for comparison was limited. The NYC Health Department continues to monitor this public health threat, and clarify factors associated with CRO acquisition, ultimately to help control CRO spread into the community.

## Background

Many carbapenem-resistant organisms (CRO), such as carbapenem-resistant Enterobacterales (CRE), are considered urgent or serious human health threats.[1] Carbapenems are often treatments of last resort for infections resistant to other antibiotics,[2] making CRO harder to treat than susceptible organisms. CRO are particularly concerning because they can harbor plasmids containing carbapenemase genes, such as *Klebsiella pneumoniae* carbapenemase (KPC) and New Delhi Metallo-β-Lactamase (NDM), which encode for enzymes that break down carbapenems. [3] Plasmids are highly transmissible between bacteria, facilitating rapid spread of resistance. Recent studies from New York City (NYC) and Canada found carbapenemase-containing plasmids in approximately 25% of CRE.[4, 5]

While CRO epidemiology has historically focused on hospital-acquired infections, there is increasing interest in community-associated incidence. [6–11] Several studies have used laboratory-based surveillance and medical record review to identify CRE cases and have categorized them as community-associated (with and without travel), healthcare-associated community onset (with and without travel), or healthcare-associated. [6, 9, 12–16] There is no definition consistently used for CRO identified in the community. [7–9]

In 2018, a NYC Health Code amendment required laboratories to report CRE isolated among NYC residents to the NYC Health Department, including antibiotic susceptibility testing results, and when available, genetic and/or phenotypic testing for the presence of a carbapenemase. Some laboratories voluntarily reported other CRO results in addition to CRE. We aimed to identify, quantify, and characterize community-associated CRO (CA-CRO) among all CRO reported in NYC, the scope of which was unknown prior to this project, through NYC laboratory-reportable surveillance data and medical record review. We pilot tested conducting patient or proxy telephone screening and interviews, since laboratory-reportable data and medical records alone may not accurately determine if a patient meets CA-CRO eligibility criteria.[6, 7, 13] As a proof-of-concept, we also aimed to explore whether genomic clustering linked to epidemiologic data could generate hypotheses about CRO transmission. Here we describe CA-CRO identified through exploratory investigations as part of routine surveillance.

## Methods

The NYC Health Department receives CRO laboratory reports through the New York State Electronic Clinical Laboratory Reporting System. Reports are automatically routed to the NYC Health Department surveillance and case management system, Maven, version 5.2.4.2 (Conduent Software, Florham Park, NJ). Our CA-CRO case definition was patients with bacterial cultures from urine or skin specimens that grew gram-negative organisms exhibiting carbapenem resistance (excluding intrinsic carbapenem resistance, such as imipenem-resistant *Morganella morganii* lacking other carbapenem resistance) among NYC residents aged ≤70 years with no international travel, and no hospitalizations or long-term care facility (LTCF) stays greater than 24 hours, within 12 months before specimen collection. If >1 specimen was received per patient, to the best of our ability we anchored our investigations on the earliest specimen collection date within the study window. We included only urine and skin cultures, since other specimen types, like blood, respiratory, and rectal specimens, are more commonly collected from in-patient hospital or LTCF settings. Similarly, we restricted patient age to ≤70 years, as younger people are less likely to have hospitalizations or LTCF stays within the past 12 months than older people. These restrictions were made to accommodate investigator capacity limitations. Additionally, we excluded patients with international travel within the past 12 months to focus on local acquisition.

To identify potential CA-CRO cases, patients with CRO cultures collected from December 2020 through May 2023 were screened for eligibility. For patients ≤70 years, with a CRO cultured from urine or skin specimens, and who met residence criteria, medical record reviews were conducted using any available information in the NYC Regional Health Information Organizations (RHIOs), Healthix and BronxRHIO. [17–20] Telephone calls with patients or proxies enabled additional screening. Potentially eligible patients who were not reached, had no available proxy, or refused screening/interview were excluded.

During screening or interview phone calls, patients confirmed or provided demographic information. Missing/unknown demographic variables were updated from Maven, if available. Through interviews, we explored the epidemiologic utility of obtaining richer medical and behavioral histories than are available in medical records. We asked patients with a CA-CRO additional questions organized into three domains: comorbidities at the time of specimen collection; other clinical and medical history prior to specimen collection (e.g. indwelling device use [e.g. urinary catheter, ventilator, feeding tube]); and behavioral history prior to specimen collection (e.g. sexual and reproductive history, drug use, and animal exposures). Three interview questions added during data collection were not asked of all patients (history of an outpatient visit, outpatient procedure, and pregnancy in the year prior to specimen collection). Responses to medical and behavioral history questions that were missing/unknown for any reason were excluded from denominators. Interview responses were collected in REDCap 13.1.30.

To contextualize patients with a CA-CRO, we conducted a descriptive comparison of available demographics for NYC using the 2016–2020 Integrated Public Use Microdata Series data, restricting to the population aged ≤70 years.[21] We used publicly available data from the annual cross-sectional 2019 and 2008 Community Health Survey (CHS), to estimate the prevalence of available health behaviors and comorbidities among NYC residents.[22, 23] These survey years include the most recent iterations of a limited number of questions comparable to our interview questions. The CHS populations of non-institutionalized adults aged 18–74 years in the 2019 survey and 18–64 in the 2008 survey most closely matched our study population, and estimates were weighted to the NYC population.[23] To assess health inequities, area-based poverty using patient ZIP Code was defined as the percent of residents with incomes below the federal poverty level, per American Community Survey (ACS), 2018–2022.[24]

Once we identified patients with a CA-CRO, we contacted the NYC Public Health Laboratory (PHL) and the state public health laboratory, Wadsworth Center (WC), to request whole genome sequencing (WGS) on available isolates from those patients. PHL and WC retain a repository of submitted CRO specimens and records of testing results. Clinical laboratories may submit CRO specimens to PHL or WC for additional testing if they lack the ability to perform molecular and/or phenotypic carbapenemase testing themselves or for additional testing requested by PHL, WC, or the United States (U.S.) Centers for Disease Control and Prevention (CDC) based on initially reported results. WGS was performed using Illumina-based technologies.[25]

Two types of cluster analyses were conducted. WC compared sequenced isolates from patients with a CA-CRO with a reference database that included isolates from these investigations and isolates sequenced at WC between 2017–2023. This WC relatedness analysis compared isolates of the same multi-locus sequence type (MLST). When available, the number of mutation events (MEs) separating each isolate from its nearest neighbor was reported, with MEs defined as short insertions or deletions or single nucleotide polymorphisms (SNPs).[26] The second analysis, conducted by PHL, used the National Center for Biotechnology Information (NCBI) Pathogen Detection tool to examine potential clusters of isolates sequenced from these investigations with other sequences uploaded in NCBI.[27]

Descriptive data summaries were compiled in SAS Enterprise Guide 7.15 (SAS Institute, Cary, NC). These activities were considered public health surveillance and deemed exempt from review by the NYC Health Department Institutional Review Board. See Supplemental Materials for additional details on methods.

## Results

During December 2020–May 2023, 6,563 CRO isolates were collected from 3,708 NYC patients, and reported to the NYC Health Department. We identified 379 patients (10% of 3,708) for medical record review based on initial laboratory report screening, and 36 of these met our case definition (Figure 1). Of note, 39 patients who remained eligible after medical record review could not be reached for telephone screening to confirm their CA-CRO eligibility. The interviewed patients, those screened and excluded, and those unresponsive to screening appeared dissimilar across demographic variables (Supplemental Table 1). For instance, compared to the other groups, cases tended to be older and those who were unresponsive to screening were more often female and living in low poverty areas, and less often Latino.

**Figure 1.**
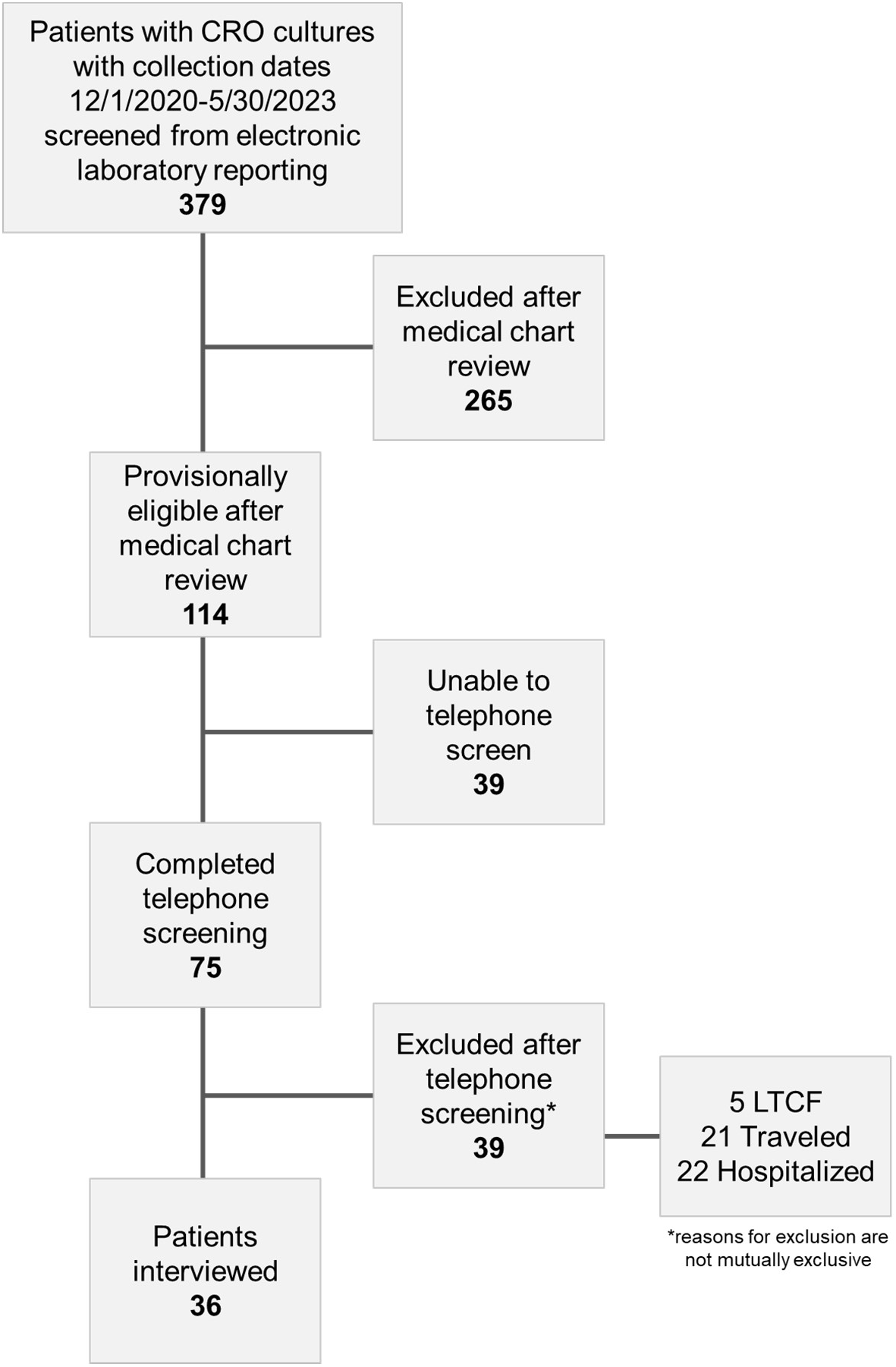
Flow diagram for patient inclusion* into Community-Associated Carbapenem Resistant Organism surveillance investigations among New York City residents, December 2020–May 2023 Abbreviations: CRO = carbapenem-resistant organism, LTCF = long-term care facility *CA-CRO cases were defined as gram-negative bacterial cultures, from urine or skin specimens, exhibiting carbapenem resistance, among New York City residents aged ≤70 years with no international travel and no hospitalization or long-term care facility stays greater than 24 hours within 12 months before specimen collection.

For the 36 patients meeting the case definition, 34 interviews were conducted with the patient and 2 with the patients’ proxy. The median time to interview was 23 days (interquartile range [IQR]: 15–96 days). Patient demographic characteristics are shown in Table 1; 61% were female; 39% were Latino, 19% Black, and 19% white; and the median age was 61 years (IQR: 45–68 years). Gender was concordant with sex at birth for all patients with a CA-CRO. The median percent of the population living below 100% of the federal poverty level in the ZIP codes where patients with a CA-CRO lived was 17% (IQR: 11%–24%). In our exploratory comparison with NYC residents 70 years or younger, patients in this investigation were disproportionately female (61% in the current investigation vs. 51% in NYC), Latino (39% vs. 30%), non-white (81% vs. 69%), 65–70 years old (36% vs. 6%), and born outside of the U.S. (50% vs. 37%), and lived in ZIP codes with similar area-based poverty (17% vs 12%).

**Table 1.** Demographics of New York City residents meeting the Community-Associated Carbapenem Resistant Organism (CA-CRO) case definition,* December 2020–May 2023.

| Demographic characteristics† | Patients with a CA-CRO*<br>(N=36) | New York City‡ |
| --- | --- | --- |
| Age at time of specimen collection in years<br>(median [interquartile range]) | 61 (45, 68) | 33 (19, 50) |
| Age category (years) |  |  |
| <18 | 0 (0%) | 23% |
| 18-24 | 0 (0%) | 9% |
| 25-44 | 9 (25%) | 35% |
| 45-64 | 14 (39%) | 27% |
| 65-70 | 13 (36%) | 6% |
| Sex and Gender |  |  |
| Female | 22 (61%) | 51% |
| Male | 14 (39%) | 49% |
| Sexual Orientation |  |  |
| Heterosexual | 31 (86%) | Unavailable |
| Gay, Lesbian or Bisexual | 1 (3%) |  |
| Declined or Missing | 4 (11%) |  |
| Born outside of the United States |  |  |
| Yes | 18 (50%) | 37% |
| No | 14 (39%) | 63% |
| Declined or Missing | 4 (11%) | 0% |
| Race and Ethnicity§ |  |  |
| Latino | 15 (42%) | 30% |
| Asian | 4 (11%) | 15% |
| Black or African American | 7 (19%) | 22% |
| White | 8 (22%) | 31% |
| Other | 1 (3%) | 4% |
| Does not identify with any race | 1 (3%) | 0% |
| Declined or Missing | 0 | 0% |

| Demographic characteristics<sup>†</sup> | Patients with a CA-CRO*<br>(N=36) | New York City<sup>‡</sup> |
| --- | --- | --- |
| **Employment Status** |  |  |
| Employed | 11 (31%) | 52% |
| Out of workforce<sup>¶</sup> | 14 (39%) | 24% |
| Unemployed | 7 (19%) | 4% |
| Declined or Missing | 4 (11%) | 20% |
| **Borough<sup>¶</sup>** |  |  |
| Bronx | 6 (17%) | 17% |
| Brooklyn | 6 (17%) | 31% |
| Manhattan | 9 (25%) | 19% |
| Queens | 14 (39%) | 27% |
| Staten Island | 1 (3%) | 6% |
| **Percent of residents in patient's ZIP code below 100% of the federal poverty limit (median [interquartile range])<sup>‡</sup>** | 17% (11%, 24%) | 12% (9%, 20%) |
| **Neighborhood poverty<sup>‡</sup>** |  |  |
| 0 to <10% (low poverty areas) | 6 (17%) | 22% |
| 10 to <20% (medium poverty areas) | 17 (47%) | 47% |
| 20 to <30% (high poverty areas) | 9 (25%) | 20% |
| 30 to 100% (very high poverty areas) | 4 (11%) | 12% |

\*CA-CRO cases were defined as gram-negative bacterial cultures, from urine or skin specimens, exhibiting carbapenem resistance, among New York City residents aged ≤70 years with no international travel and no hospitalization or long-term care facility stays greater than 24 hours within 12 months before specimen collection.
<sup>†</sup>N (%) except where noted.
<sup>‡</sup>Available variables were summarized for comparison with New York City from the American Community Survey 2018-2022, all ages, for neighborhood poverty measures, and Integrated Public Use Microdata Series data 2016-2020, limited to New York City residents aged 70 years and younger, for all other comparisons.
<sup>§</sup>Race and ethnicity were collapsed into one category and Latino is Hispanic or Latino of any race. All other patients were categorized by their race. Declined or missing was used for patients missing both their race and ethnicity.
<sup>¶</sup>Out of workforce includes individuals who are retired, disabled, students, or stay-at-home parents
<sup>¶</sup>In New York City, each of the five boroughs are coterminous with a county: the Bronx is Bronx County; Brooklyn is Kings County; Manhattan is New York County; Queens is Queens County, and Staten Island is Richmond County.

Table 2 includes medical and behavioral history variables representing each of the three question domains; Supplemental Table 2 provides additional history data. Over half of patients with a CA-CRO had at least one comorbidity and approximately a quarter had two or more; in particular, 25% had diabetes. Twenty-two percent of patients reported a condition that limited their mobility and/or used an assistive device such as a wheelchair, walker, or cane. In the 12 months before specimen collection, 79% of patients had ≥1 outpatient appointment, 56% had ≥1 outpatient surgery or procedure; and 33% reported receiving homecare from a nurse, doctor, or aide. Within three months before specimen collection 38% of patients reported taking antibiotics; within two days before specimen collection 25% of patients had a urinary catheter or indwelling device. Forty-one percent of patients were sexually active, 22% reported contraceptive use in the prior year, and none reported a recent diagnosis of a sexually transmitted infection or injection drug use. Within the 3 months prior to specimen collection, 27% of patients reported ≥1 encounter with a live animal including dogs, cats, and birds.

**Table 2.** Self-reported medical and behavioral histories among New York City residents meeting the Community-Associated Carbapenem Resistant Organism (CA-CRO) case definition,* December 2020–May 2023.

| Domains and questions | Patients with a CA-CRO<br>no. responded/total (%)<br>n = 36 |
| --- | --- |
| **Comorbidities at the time of specimen collection** |  |
| Any comorbidity<sup>†</sup> | 21/36 (58%) |
| Number of comorbidities |  |
| 0 | 15/36 (42%) |
| 1 | 13/36 (36%) |
| 2+ | 8/36 (22%) |
| Diabetes | 9/36 (25%) |
| **Other clinical and medical history prior to specimen collection** |  |
| Outpatient appointment in the year prior | 19/24 (79%) |
| Outpatient surgeries or procedures in the year prior | 15/27 (56%) |
| Homecare from a nurse, doctor, or aide in the year prior | 12/36 (33%) |
| Antibiotic use in the 3 months prior | 12/32 (38%) |
| Indwelling catheters or devices within 2 calendar days | 9/36 (25%) |
| Limited ambulation or the use of a wheelchair, walker, cane, etc. | 8/36 (22%) |
| Non-surgical wound in the prior year | 8/36 (22%) |
| Household member spent at least 1 night in a hospital in prior year | 4/36 (11%) |
| Sexually transmitted infection diagnosis prior year | 0/29 (0%) |
| **Behavioral history prior to specimen collection** |  |
| Sexual contact within prior year | 13/32 (41%) |
| Use of contraceptives (self or partner) within prior year | 6/27 (22%) |
| Encounters with live animals within the prior 3 months | 9/33 (27%) |
| Injection drug use prior 90 days | 0/33 (0%) |

\*CA-CRO cases were defined as gram-negative bacterial cultures, from urine or skin specimens, exhibiting carbapenem resistance, among New York City residents aged ≤70 years with no international travel and no hospitalization or long-term care facility stays greater than 24 hours within 12 months before specimen collection.
<sup>†</sup>Any comorbidity is defined as any cancer, diabetes (type 1 or 2), lung disease (asthma, chronic obstructive pulmonary disease, chronic bronchitis, emphysema, cystic fibrosis), heart disease (coronary artery disease [heart attack, congestive heart failure, stroke], congenital heart disease), gastrointestinal or hepatic disease (cirrhosis, chronic liver diseases), immunodeficiency or immunosuppression, HIV, organ transplant, or renal failure with dialysis.

Compared with CHS, a higher percentage of patients with a CA-CRO reported ever being told by a health professional that they had diabetes (25% vs. 11% in CHS) and reported using any assistive devices (22% vs. 7%, including aids to ambulation and hearing-assistive telephone in CHS). Patients with a CA-CRO were less likely to report sexual contact within the prior year (41% vs. 75%), and less likely to have used a form of contraception (22% vs. 56%) during sexual activity. CHS estimates were similar to those of patients with a CA-CRO who reported an outpatient appointment in the past year (79% vs. 85% who had seen any doctor, nurse, or other health professional in the last 12 months in CHS), who reported lung disease (14% vs. 16%, excluding cystic fibrosis in CHS), and who reported a live animal encounter within the prior 3 months (27% vs. 28%, living in a household with dogs or cats in CHS).

Most isolates were cultured from urine (94%), and the remainder were from skin (6%). Nearly 70% were either *Escherichia coli* (39%) or *Klebsiella pneumoniae* (28%), and the remaining isolates included *Enterobacter cloacae* complex, *Klebsiella aerogenes*, *Pseudomonas aeruginosa*, *Proteus mirabilis*, and *Pluralibacter gergoviae* (Figure 2). Most CA-CRO isolates (53%) did not undergo carbapenemase testing and were not available for further characterization. Among those tested, KPC and NDM were detected in 41% and 6% of isolates, respectively. No carbapenemase gene was detected using molecular or phenotypic tests in the remaining isolates tested (53%).

**Figure 2.**
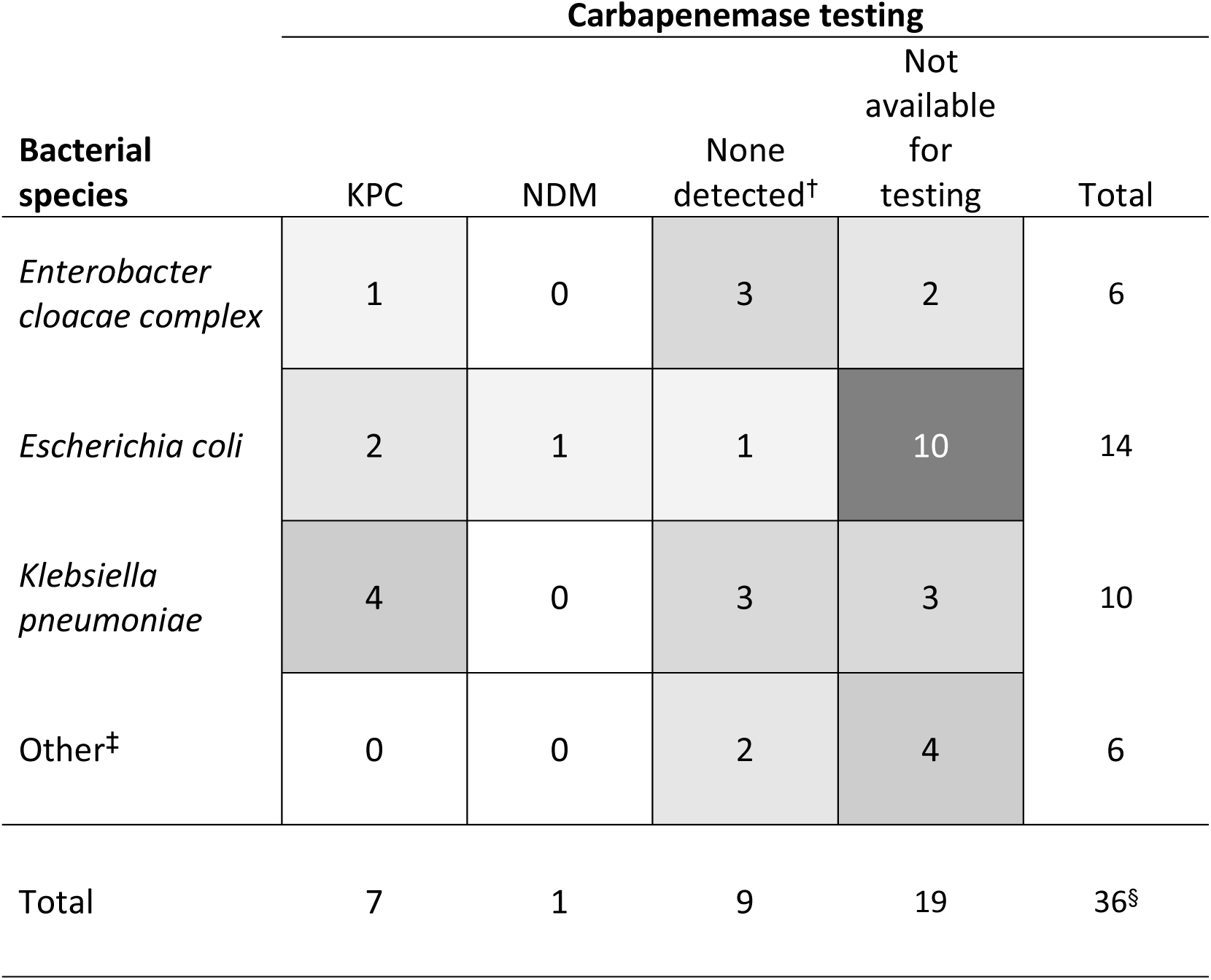
Heat map of bacterial species by carbapenemase testing status for Community-Associated Carbapenem Resistant Organism (CA-CRO) cases* among New York City residents, December 2020–May 2023 Abbreviations: KPC = Klebsiella pneumoniae Carbapenemase, NDM = New Delhi metallo beta-lactamase *CA-CRO cases were defined as gram-negative bacterial cultures, from urine or skin specimens, exhibiting carbapenem resistance, among New York City residents aged ≤70 years with no international travel and no hospitalization or long-term care facility stays greater than 24 hours within 12 months before specimen collection. †No carbapenemase detected using molecular or phenotypic tests ‡Other includes Klebsiella aerogenes, Pseudomonas aeruginosa, Proteus mirabilis, and Pluralibacter gergoviae ^§^Only one specimen per CA-CRO patient was included here. If >1 specimen was received per patient, to the best of our ability we anchored our investigations on the earliest specimen collection date within the study window.

Roughly one third of patients with a CA-CRO had a bacterial isolate available for WGS; specifically, 15 isolates from 13 patients were sequenced (Supplemental Table 3). Nine isolates with MLST matches in the WC reference database underwent the relatedness analysis. The nearest neighbors ranged from 0 MEs apart (NY-NYCPHL-CR0000000004 and NY-NYCPHL-CR0000000005, from the same patient collected one month apart) to 3147 MEs apart (NY-NYCPHL-CR0000000010). Two isolates were from the same patient (NY-NYCPHL-CR0000000006 and 2023HL-00510, *K. pneumoniae*), collected ten months apart, and were separated by 1727 MEs. This high ME separation is greater than would be predicted by the expected rate of evolution.[28] We cannot exclude the possibility that recombination contributed to this large ME separation; however, the isolates could also represent distinct infections with different bacterial strains in the same patient.[29–31] In contrast, an isolate from 2021 (NY-NYCPHL-CR0000000002, *K. pneumoniae*) was 11 MEs from a previously sequenced 2018 isolate in the reference database from the same patient, consistent with the expected rate of evolution.[28]

The Pathogen Detection BioProject ID for this analysis is PRJNA1106484. One of the 15 isolates (NY-NYCPHL-CR0000000011) failed Pathogen Detection’s quality control for lacking sufficient numbers of identified whole-genome MLST loci and was excluded. Nine isolates did not form clusters with other submitted sequences on NCBI. As of January 30, 2025, the remaining five isolates formed clusters with isolates in NCBI and were assigned SNP Cluster IDs. Isolates NY-NYCPHL-CR0000000004 and NY-NYCPHL-CR0000000005 (both *K. pneumoniae*) from the same patient one month apart formed a 2-isolate SNP cluster, PDS000201537. Isolate NY-NYCPHL-CR0000000002 (*K. pneumoniae*) was a member of the 2-isolate SNP cluster PDS000187665, which included the isolate from the same patient from 2018. Isolate NY-NYCPHL-CR0000000007 (*K. pneumoniae*) was a member of SNP cluster PDS000050967 which had 78 isolates, all from the U.S. Isolate 2022HL-01891 (*E. coli)* was a member of SNP cluster PDS000138448 which included 53 isolates predominantly from the U.S., though some were from Canada, France, and Australia.

## Discussion

Patients with a presumed CA-CRO represented a small fraction of all patients with CRO reported in NYC during the study period. Despite having no recent hospitalization or LTCF residence, patients with a CA-CRO were of an older age and medically complex. Most had at least one comorbidity, over half had a recent outpatient medical procedure, one third accessed home health care services, one-quarter had urinary catheters or indwelling devices, and nearly one-quarter had limited ambulation. Recent antibiotic use was also commonly self-reported.

Comparisons to citywide estimates, when available, contextualized findings from medical and behavioral interview questions. Patients with a CA-CRO had higher percentages of diabetes and limited ambulation, and lower percentages of sexual activity and contraception use, compared with NYC residents, which may correlate with the overall older age of patients with a CA-CRO. Additionally, CA-CRO were disproportionately detected among those who were aged 65–70 years, female, born outside of the U.S., and Latino. Race, ethnicity and socioeconomic status have been associated with increased colonization or infection with antibiotic resistant pathogens, which may be related to inadequate health insurance coverage or healthcare access, leading to delays in care and inappropriate or incomplete treatment.[32, 33] More work is needed to identify underlying causes, since many socioeconomic factors are collinear.[32]

Our epidemiologic findings are consistent with other CA-CRO reports, with respect to higher median age, and higher proportions of female patients and patients with comorbidities.[7, 9, 12, 13, 16] Approximately ten percent of patients in our population had cared for family members with recent hospitalization, similar to findings from a Colorado study from 2014–2016, among patients with a CA-CRE.[7] KPC was the most common carbapenemase identified among our cases, similar to findings in other studies that focused on CA-CRE.[14, 16]

For the first time in NYC, we examined CA-CRO isolates in the context of their genetic relationships with other sequenced CRO. Results from the two WGS analyses were generally concordant. For most patients with a CA-CRO no isolate was available for WGS, and genomic clustering occurred in fewer than half of sequenced isolates. We suspect this reflects a relatively low volume of CRO being sequenced and shared to public repositories, offering only superficial coverage of the circulating CRO isolates across all setting types; however, we cannot rule out the possibility that the limited clustering signals novel CRO strains. Antimicrobial Resistance Laboratory Network (ARLN) guidance now recommends WGS for certain CRO to better identify and respond to new or emerging antibiotic-resistant threats.[34]

Increasingly, genomic data can help identify clusters and illuminate underlying contact networks that have led to transmission.[35–38] In hospital settings, WGS has been used for antimicrobial-resistant bacteria surveillance to detect nosocomial transmission and target rapid interventions.[39] Although challenging, WGS can also be used to identify clusters of resistant bacteria in community settings. For instance, Pathogen Detection was used to investigate a Massachusetts CRO cluster where the patients had no obvious healthcare links and where human isolates matched closely with companion animal and environmental isolates from a veterinary hospital CRO outbreak. Genomic evidence supported the hypothesis that transmission to humans likely occurred via contact with companion animals or the veterinary hospital environment.[40]

In our investigation, we explored the utility of leveraging available epidemiologic data integrated with sequence data. The public health surveillance databases contain metadata that can be linked to each sequenced isolate including ancillary specimen, patient, and laboratory data. Using these metadata, we were able to describe many of the relationships identified in the relatedness analysis (e.g., when related isolates were from prior specimens cultured from the same individuals). In contrast, describing the epidemiologic relationships using NCBI metadata alone was less practical, since only higher-level and de-identified metadata were available—such as the state of origin, host species, and specimen collection year. Potential exists for WGS to suggest how transmission occurred, illustrated by the example above. However, because of low CRO sequencing coverage and sparse metadata, we were unable to realize that potential.

One strength of this investigation was the rich data gathered through interviews, and telephone screening allowed us to exclude 39 patients with a CRO whose infections would have been considered community-associated, according to our case definition, by medical record review alone. Another strength was the development of processes with our laboratory colleagues to incorporate genomic analyses with epidemiologic surveillance. Additionally, mandated local reporting of CRE to the NYC Health Department provided high-volume CRE data for a large urban jurisdiction, which facilitated identification of CA-CRO as a currently uncommon but still concerning condition. However, since only CRE reporting was mandated during the study period, non-Enterobacterales CRO were underrepresented. The Health Code was amended in October 2023 to mandate reporting of all CRO.[41]

Our analysis was limited by screening non-response and missing data; patients unable to be screened had different demographics than patients with a CA-CRO, and results may not be generalizable to all patients with a CA-CRO in NYC. Interview questions relied on self-reported data, and the NYC Health Department COVID-19 pandemic response limited staff capacity to consistently conduct timely interviews, both of which could have contributed to recall bias. We lacked a comparison population with all the measures we included here. The CHS collected some similar measures, but interview questions were not always asked in the same way, and CHS does not cover certain groups (e.g., adults in households with no telephone service or living in LTCF).[23]. That said, these exclusions may have made the CHS population more similar to patients with a CA-CRO in our investigations, who also were required to have telephone service to participate in the screening and interview calls, and to reside outside of LTCF.

Clinical laboratories often have limited specimen retention capacity, resulting in a short time window to request specimen submission for additional testing, including carbapenemase detection and WGS. Isolates available for comparison in both WGS analyses were limited; neither likely represent the true diversity of circulating CRO. Similarly, since colonization screening cultures seldom occur outside of hospitals and LTCFs, and reportable laboratory cultures are primarily triggered by symptomatic infections, surveillance data are unlikely to represent the true community CRO prevalence. Furthermore, patients could have acquired CRO in an inpatient setting, remaining colonized >12 months without culture, and our definition would characterize them as CA-CRO at the time of specimen collection. At the time of these investigations, the nationally accepted CRO case definition considered each case as unique if it was ≥1 year after any previous case in the same patient of the same species/carbapenemase combination; currently each species/carbapenemase combination detection is considered lifelong.[42, 43] Given this revision, some CA-CRO cases may have been misattributed to this investigation’s time frame or misclassified as CA-CRO instead of hospital-acquired CRO. Additionally, selecting appropriate thresholds of continuous variables (MEs or SNPs) to define clusters of related isolates is an area of ongoing debate.[44]

Comparison of these results to other CA-CRO studies is limited, because, although many of the elements that define CRO in the community are conserved across studies, we lack a standard CA-CRO case definition.[8]

## Conclusion

As defined, CA-CRO was uncommonly diagnosed through May 2023. However, the NYC Health Department will continue to monitor CA-CRO incidence. PHL is actively planning to expand its sequencing capacity to routinely sequence carbapenemase-producing CRO isolates, in accordance with ARLN recommendations.

Future CA-CRO study would benefit from a standard case definition and the identification of an appropriate comparison group with more concordant epidemiological data. A process for investigating antibiotic resistant organisms that securely shares genomic and epidemiologic information from public health jurisdictions nationwide, such as CDC’s System for Enteric Disease Response, Investigation, and Coordination, and Tuberculosis Genotyping Information Management System[36, 45], could allow for generating transmission-related hypotheses outside of inpatient healthcare settings.

## Data Availability

Data from this manuscript is not available.

## Contributions

Conception and design of the work: KD, MK, CS

Acquisition of data: DB, KD, TP, CS, YL, USK

Analysis of data: TP, CS, RZ, CP, SA, FT, KC

Interpretation of data: all

Drafting the manuscript: CS, RZ, KAA, NB, SA, CP, FT, WGG, MK

Reviewing the manuscript and final approval: all

## Acknowledgements

We gratefully acknowledge the comments and advice of Scott Harper, Sharon Greene, Kenya Murray (currently at the University of Georgia), Rachel Levit, and Don Weiss from the NYC Health Department Bureau of Communicable Diseases as well as Nina Grossman, Aaron Olsen, Rain J. Wiegartner, and Jorge Montfort Gardeazabal from the NYC Public Health Laboratory. We thank the Wadsworth Center advanced Genomic Technologies Center for conducting sequencing. An earlier version of the findings from these investigations was presented at the SHEA Spring conference 2024.

