## Supplemental materials for "Community-associated Carbapenem-Resistant Organism Case Investigations in New York City"

**Affiliations:**
[1]Bureau of Communicable Diseases, New York City Department of Health and Mental Hygiene,
[2]Department of Epidemiology and Biostatistics, City University of New York Graduate School of Public Health and Health Policy,
[3]Public Health Laboratory, New York City Department of Health and Mental Hygiene,
[4]Wadsworth Center, New York State Department of Health

### Methods

*Interview Details*

Patient interviews were conducted in the preferred language of the patient and if needed an interpreter service was used. Any references to patient screening or patient interviews encompass screening or interviews conducted with patients or their proxies (if the patient was unable to speak on the phone); all questions asked to proxies were in relation to the patient of interest.

*Demographic variables*

Patients were asked about both sex at birth and gender. We defined a single race and ethnicity variable, where the Latino category included Hispanic or Latino ethnicity of any race, and other categories included only patients of non-Hispanic or Latino ethnicity. Employment was defined based on patients' self-reported occupation. We calculated patient age at specimen collection using patients' birth date and specimen collection date.

*Questionnaire details*

Comorbidities included cancer, diabetes (type I or II), lung disease (asthma, chronic obstructive pulmonary disease, chronic bronchitis, emphysema, cystic fibrosis), heart disease (coronary artery disease [heart attack, congestive heart failure, stroke], congenital heart disease), gastrointestinal or hepatic disease (cirrhosis, chronic liver diseases), immunodeficiency or immunosuppression, HIV, organ transplant, or renal failure with dialysis. Animal exposures were defined broadly as exposures to any animals (which could include companion animals, contact with pets belonging to people in patient's social networks, visits to or working at a farm or zoo, among other possibilities). From February 2021 through September 2021 three questions were added—outpatient appointments, outpatient procedures, and pregnancy within the year prior to specimen collection. No additional questions were added from October 2021 through the end of the interview period in June 2023.

*Screening non-response*

Basic demographic variables available from Maven or telephone screening were used to describe all patients provisionally eligible after medical record review by final eligibility status. For demographics reported, responses to the demographic questions from screening or interviews were prioritized for reporting. However, if any demographic variable was unknown or missing following screening/interview, available data in the Maven surveillance system was used to supplement this information. Limited demographic information was available in the Maven surveillance system and only data from Maven was available for patients who were not able to be screened, which determined the variables for comparison of these three patient populations.

*Comparison variables from Community Health Survey*

There were a limited number of variables from the Community Health Survey (CHS), which uses stratified random sampling, that were comparable to the questions from patient interviews. From the 2008 CHS, we used *owndogcat* ("Do you or anyone else in the household have any dogs or cats?"). From the 2019 CHS, we used *diabetes19* ("Have you ever been told by a doctor, nurse or other health professional that you have diabetes?"), *assistdevice* ("Do you use any assistive devices, such as a cane, a wheelchair, an adapted bed, or a hearing assistive telephone because of a health condition?”), *visitnonpcp12m19* (“Have you seen any doctor, nurse, or other health professional in the last 12 months?”), *sexuallyactive19* (“Sexually active in past 12 months”), and *bthcontrollastsex19* (“Any birth control at last sex (includes condom use)”).

*Whole-genome sequencing methods*

At Wadsworth Center (WC), genomic DNA was extracted on the QIAcubeHT and sequencing was performed on the Illumina NextSeq as previously described. [1] MLST and AR gene identification was performed using an in-house developed bioinformatic pipeline. [2] A relatedness analysis using a separate in-house developed bioinformatic pipeline [3] was conducted to compare each isolate to other isolates previously sequenced at WC from the same species and MLST. Briefly, the pipeline clusters similar isolates using k-mer-based analysis, then uses an internal reference genome within each cluster for reference-based alignment and calling of mutation events (MEs), consisting of SNPs and short insertion/deletion events. As a note, WC serves multiple jurisdictions in the northeast region and specimens in the reference database for comparison in the relatedness analysis represent specimens from these jurisdictions.

For the whole-genome sequencing of CA-CRO at New York City Public Health Laboratory (NYC PHL), the DNA extracts were subjected to a tagmentation-based library preparation using the Illumina DNA Prep Kit (Cat. # 20018705) with IDT for Illumina Unique Dual Index Sets (Cat. #s: 20027213(UD-A), 20027214(UD-B), 20042666(UD-C), 20042667(UD-D)). The libraries were run on an Illumina MiSeq at 251bp paired-end reads with a llumina MiSeq Reagent Kit 600-cycle v3 kit (Cat. # MS-102-3003) or a NextSeq2000 with a NextSeq 1000/2000 P2 Reagent 300-cycle kit v3 kit (Cat. # 20046813) at 151 bp paired end reads. The pathogen genome sequence files (FASTQ) and associated metadata were submitted to the NCBI Pathogen Detection [4] (results accessed January 30, 2025) to identify potential clusters with other submitted sequences.

Pathogen Detection uses two clustering pipelines to identify genomic relationships for submitted sequence. The first pipeline uses a reference wgMLST scheme to identify the loci and alleles in each assembled genome. A 25-allele cut-off is used to cluster related isolates. The second pipeline initially uses k-mer distances to cluster related isolates which follows by a SNP analysis. 50-SNP single-linkage clustering is used to create the clusters. The wgMLST method is used for the organisms with large numbers of isolates (≥1,000 isolates), while the second pipeline is used for those organisms with that have less than 1,000 isolates.

### Results

*Screening non-response*

Compared to patients with a CA-CRO, patients screened and excluded tended to be younger; more evenly distributed with regard to sex; less likely to be white; more likely to live in the Bronx and Brooklyn and less likely to live in Manhattan or Queens; and less likely to live in low poverty areas and more likely to live in very high poverty areas. Compared to patients with a CA-CRO, patients unable to be screened tended to be younger, more likely to be female; less likely to be Latino and more likely to be white; less likely to live in the Bronx and Manhattan and more likely to live in Brooklyn and Staten Island; and live in lower poverty areas. See Supplemental Table 1.

Supplemental Table 1. Demographics of New York City residents meeting the Community-Associated Carbapenem Resistant Organism (CA-CRO) case definition* after medical chart review, by telephone screening and final eligibility status, December 2020–May 2023

| **Demographic characteristics**† | **Patients with a CA-CRO* (N=36)** | **Patients screened and excluded (N=39)** | **Patients unable to be screened (N=39)** |
|---|---|---|---|
| **Age at time of specimen collection in years (median [interquartile range])** | 61 (45, 68) | 52 (37, 64) | 56 (41, 63) |
| **Age category (years)** | | | |
| <18 | 0 (0%) | 2 (5%) | 2 (5%) |
| 18-24 | 0 (0%) | 1 (3%) | 2 (5%) |
| 25-44 | 9 (25%) | 10 (26%) | 8 (21%) |
| 45-64 | 14 (39%) | 20 (51%) | 19 (49%) |
| 65-70 | 13 (36%) | 6 (15%) | 8 (21%) |
| **Sex** | | | |
| Female | 22 (61%) | 22 (56%) | 33 (85%) |
| Male | 14 (39%) | 17 (44%) | 6 (15%) |
| **Race and Ethnicity**‡ | | | |
| Latino | 15 (42%) | 15 (39%) | 9 (23%) |
| Asian | 4 (11%) | 6 (15%) | 5 (13%) |

| Demographic characteristics† | Patients with a CA-CRO* (N=36) | Patients screened and excluded (N=39) | Patients unable to be screened (N=39) |
|---|---|---|---|
| Black or African American | 7 (19%) | 8 (21%) | 8 (21%) |
| White | 8 (22%) | 6 (15%) | 10 (26%) |
| Other | 1 (3%) | 1 (3%) | 0 (5%) |
| Does not identify with any race | 1 (3%) | 2 (5%) | 4 (10%) |
| Declined or Missing | 0 (%) | 0 (0%) | 3 (8%) |
| **Borough§** | | | |
| Bronx | 6 (17%) | 12 (31%) | 5 (13%) |
| Brooklyn | 6 (17%) | 12 (31%) | 9 (23%) |
| Manhattan | 9 (25%) | 5 (13%) | 7 (18%) |
| Queens | 14 (39%) | 9 (23%) | 13 (33%) |
| Staten Island | 1 (3%) | 1 (3%) | 5 (13%) |
| **Percent of residents in patient's ZIP code below 100% of the federal poverty limit (median [interquartile range])‖** | 17% (11%, 24%) | 17% (12%, 26%) | 14% (10%, 21%) |
| **Neighborhood poverty‖** | | | |
| 0 to <10% (low poverty areas) | 6 (17%) | 4 (10%) | 10 (26%) |
| 10 to <20% (medium poverty areas) | 17 (47%) | 20 (51%) | 19 (49%) |
| 20 to <30% (high poverty areas) | 9 (25%) | 9 (23%) | 7 (18%) |
| 30 to 100% (very high poverty areas) | 4 (11%) | 6 (15%) | 3 (8%) |

*CA-CRO cases were defined as gram-negative bacterial cultures, from urine or skin specimens, exhibiting carbapenem resistance, among New York City residents aged ≤70 years with no international travel and no hospitalization or long-term care facility stays greater than 24 hours within 12 months before specimen collection.

†N (%) except where noted.

‡Race and ethnicity were collapsed into one category and Latino is Hispanic or Latino of any race. All other patients were categorized by their race. Declined or missing was used for patients missing both their race and ethnicity.

§In New York City, each of the five boroughs are coterminous with a county: the Bronx is Bronx County; Brooklyn is Kings County; Manhattan is New York County; Queens is Queens County, and Staten Island is Richmond County.

‖Neighborhood poverty measures were merged from the American Community Survey 2018-2022, all ages, by ZIP code.

Supplemental Table 2. Additional self-reported medical and behavioral histories among New York City residents meeting the Community-Associated Carbapenem Resistant Organism (CA-CRO) case definition,* December 2020–May 2023

| **Domains and questions** | **Patients with a CA-CRO*<br>no. responded/total (%)<br>n = 36** |
|---|---|
| **Comorbidities at the time of specimen collection** | |
| Any comorbidity[†] | 21/36 (58%) |
| Number of comorbidities | |
| 0 | 15/36 (42%) |
| 1 | 13/36 (36%) |
| 2+ | 8/36 (22%) |
| Individual comorbidities (not mutually exclusive) | |
| Diabetes | 9/36 (25%) |
| Type I | 1/36 (3%) |
| Type II | 7/36 (19%) |
| Unspecified | 1/36 (3%) |
| Gastrointestinal or hepatic disease | 5/34 (15%) |
| Heart disease | 5/36 (14%) |
| Lung disease | 5/36 (14%) |
| Immunodeficiency or immunosuppression | 4/36 (11%) |
| Organ transplant | 3/36 (8%) |
| Cancer | 2/35 (6%) |
| HIV | 1/36 (3%) |
| Renal failure with dialysis | 1/35 (3%) |
| **Other clinical and medical history prior to specimen collection** | |
| Outpatient appointment | 19/24 (79%) |
| Number of outpatient appointments | |
| 1 | 8/24 (33%) |
| 2 | 3/24 (13%) |
| 3 | 8/24 (33%) |
| Type of outpatient appointment (not mutually exclusive) | |
| Obstetrics/gynecology visit | 7/24 (29%) |
| Annual visit | 6/24 (25%) |
| Other primary care visit | 6/24 (25%) |
| Emergency room visit | 5/24 (21%) |
| Urology visit | 3/24 (13%) |
| Dentist visit | 2/24 (8%) |
| Otolaryngology visit | 2/24 (8%) |
| Gastroenterology visit | 2/24 (8%) |
| Dermatology visit | 1/24 (4%) |

| Domains and questions | Patients with a CA-CRO* no. responded/total (%) n = 36 |
|---|---|
| Other type of visit | 4/24 (17%) |
| Outpatient surgeries or procedures | 15/27 (56%) |
| Number of outpatient surgeries or procedures | |
| 1 | 12/27 (44%) |
| 2 | 2/27 (7%) |
| 3 | 1/27 (4%) |
| Type of outpatient surgery or procedure (not mutually exclusive) | |
| Colonoscopy | 5/27 (19%) |
| Catheter insertion or removal | 2/27 (7%) |
| Bladder procedure | 2/27 (7%) |
| Dental procedure | 2/27 (7%) |
| Endoscopy | 1/27 (4%) |
| Other procedure | 7/27 (26%) |
| Homecare from a nurse, doctor, or aide in prior year | 12/36 (33%) |
| Household member spent at least 1 night in a hospital in prior year | 4/36 (11%) |
| Any history of similar symptoms or bacterial illness | 20/36 (56%) |
| Non-surgical wound in prior year | 8/36 (22%) |
| Surgical wound in prior year | 3/35 (9%) |
| Limited ambulation or the use of a wheelchair, walker, cane, etc. | 8/36 (22%) |
| Any antibiotic use in the 3 months prior | 12/32 (38%) |
| Number of antibiotic courses in the 3 months prior | |
| 1 | 6/32 (19%) |
| 2 | 4/32 (13%) |
| 3 | 2/32 (6%) |
| Indwelling devices within 2 calendar days | 9/36 (25%) |
| Number of indwelling devices | |
| 0 | 27/36 (75%) |
| 1 | 7/36 (19%) |
| 2+ | 2/36 (6%) |
| Types of indwelling devices (not mutually exclusive) | |
| Urinary catheter | 7/36 (19%) |
| Pessary | 1/36 (3%) |
| Ventilator | 1/36 (3%) |
| Colostomy bag | 1/36 (3%) |
| Feeding tube | 1/36 (3%) |
| Tracheostomy tube | 1/36 (3%) |
| Sexually transmitted infection diagnosis prior year | 0/29 (0%) |
| **Behavioral history prior to specimen collection** | |
| Sexual contact within prior year | 13/32 (41%) |

| Domains and questions | Patients with a CA-CRO* no. responded/total (%) n = 36 |
|---|---|
| Number of sexual partners (among people with sexual contact within prior year) | |
| 1 | 10/13 (77%) |
| 2 | 1/13 (8%) |
| Use of contraceptives (self or partner) within prior year | 6/27 (22%) |
| Pregnancy (self) within prior year | 1/17 (6%) |
| Encounters with live animals within prior 3 months | 9/33 (27%) |
| Type of live animal encounter within prior 3 months (not mutually exclusive) | |
| Dogs | 7/33 (21%) |
| Cats | 3/33 (9%) |
| Birds | 1/33 (3%) |
| Travel outside of New York City prior year | 14/36 (39%) |
| Smoked cigarettes | 3/33 (9%) |
| Smoked marijuana prior 90 days | 4/33 (12%) |
| Injection drug use prior 90 days | 0/33 (0%) |

*CA-CRO cases were defined as gram-negative bacterial cultures, from urine or skin specimens, exhibiting carbapenem resistance, among New York City residents aged ≤70 years with no international travel and no hospitalization or long-term care facility stays greater than 24 hours within 12 months before specimen collection.

†Any comorbidity is defined as any cancer, diabetes (type 1 or 2), lung disease (asthma, chronic obstructive pulmonary disease, chronic bronchitis, emphysema, cystic fibrosis), heart disease (coronary artery disease [heart attack, congestive heart failure, stroke], congenital heart disease), gastrointestinal or hepatic disease (cirrhosis, chronic liver diseases), immunodeficiency or immunosuppression, HIV, organ transplant, or renal failure with dialysis.

Supplemental Table 3: Results of Relatedness Analysis* and Pathogen Detection† using the Genome Sequences from Patients with a Community-Associated Carbapenem Resistant Organism

| **WGS ID (Biosample ID)** | **Organism (Collection Year)** | **MLST‡ Number and Relatedness analysis** | **Pathogen detection BioProject ID PRJNA1106484 SNP cluster ID (# of isolates in the cluster)** |
|---|---|---|---|
| NY-NYCPHL-CR0000000003 (SAMN41839125) | *Enterobacter cloacae* (2021) | MLST ST 66<br>No MLST matches in WC database | |
| NY-NYCPHL-CR0000000009 (SAMN41839131) | *Enterobacter cloacae* (2022) | MLST ST 171<br>Nearest neighbor 212 MEs | |
| NY-NYCPHL-CR0000000011 | *Enterobacter cloacae* complex (2022) | MLST ST 40<br>No MLST matches in WC database | Failed Pathogen Detection Quality Control |
| NY-NYCPHL-CR0000000001 (SAMN41839123) | *Escherichia coli* (2020) | MLST ST 68/14<br>No MLST matches in WC database | |
| NY-NYCPHL-CR0000000010 (SAMN41839132) | *Escherichia coli* (2022) | MLST ST 73/4<br>Nearest neighbor 3147 MEs | |
| 2022HL-01891 (SAMN31854212) | *Escherichia coli* (2022) | MLST ST 410/471<br>Nearest neighbor 76 MEs | PDS000138448 (53)<br>Minimum SNP difference 35 |
| NY-NYCPHL-CR0000000002 (SAMN41839124) | *Klebsiella pneumoniae* (2021) | MLST ST 258<br>Nearest neighbor 11 MEs (same patient, specimen collected in 2018) | PDS000187665 (2)<br>Minimum SNP difference 3 |
| NY-NYCPHL-CR0000000004 (SAMN41839126) | *Klebsiella pneumoniae* (2021) | MLST ST 512<br>Nearest neighbor is NY-NYCPHL-CR0000000005 separated by 0 MEs (same patient, specimen collected one month later) | PDS000201537 (2)<br>Minimum SNP difference 0 |
| NY-NYCPHL-CR0000000005 (SAMN44347422) | *Klebsiella pneumoniae* (2021) | MLST ST 512<br>Nearest neighbor is NY-NYCPHL-CR0000000004 separated by 0 MEs (same patient, specimen collected one month earlier) | PDS000201537 (2)<br>Minimum SNP difference 0 |
| NY-NYCPHL-CR0000000006 (SAMN41839128) | *Klebsiella pneumoniae* (2022) | MLST ST 258<br>Nearest neighbor is 106 MEs; also separated by 1727 MEs from isolate 2023HL-00510 from the same patient 10 months prior | |

| WGS ID<br>(Biosample ID) | Organism<br>(Collection Year) | MLST[‡] Number and<br>Relatedness analysis | Pathogen detection BioProject ID PRJNA1106484<br>SNP cluster ID<br>(# of isolates in the cluster) |
|---|---|---|---|
| NY-NYCPHL-CR0000000007<br>(SAMN41839129) | *Klebsiella pneumoniae*<br>(2022) | MLST ST 258<br>Nearest neighbor 53 MEs | PDS000050967 (78)<br>Minimum SNP difference 17 |
| 2023HL-00510<br>(SAMN36356761) | *Klebsiella pneumoniae*<br>(2021) | MLST ST 258<br>Nearest neighbor 112 MEs; also separated by 1727 MEs from isolate NY-NYCPHL-CR0000000006 from the same patient 10 months later | |
| 2023HL-00512<br>(SAMN36356763) | *Klebsiella pneumoniae*<br>(2022) | MLST ST 1552<br>No MLST matches in WC database | |
| 2024HL-01025<br>(SAMN43802855) | *Klebsiella pneumoniae*<br>(2023) | MLST ST 133<br>No MLST matches in WC database | |
| 2023HL-00511<br>(SAMN36356762) | *Pluralibacter gergoviae*<br>(2022) | No MLST scheme available for this organism | |

[*]Isolates within 50 mutation events (MEs) of each other were considered closely related, with MEs defined as short insertions or deletions or single nucleotide polymorphisms (SNPs).

†Comparisons with >1000 isolates have a 25-allele cut-off to define a related cluster; comparisons with <1000 isolates use 50-SNP single linkage clustering to define clusters.

‡Multilocus sequence typing (MLST) is a whole genome sequencing-based laboratory method that assigns a numeric 'sequence type' based on bacterial species-specific typing schemes as a large-scale measure of genetic relatedness between strains.
